# Examining the burden of opioid prescribing for non-cancer pain considering socio-economic differences, in Wales. A retrospective database study examining trends between 2005 – 2015

**DOI:** 10.1101/19012625

**Authors:** Emma Davies, Bernadette Sewell, Mari Jones, Ceri Phillips, Jaynie Rance

**Affiliations:** College of Human and Health Sciences, Swansea University, Singleton Park, Sketty Lane, Swansea, Wales, UK, SA2 8PP; Swansea Centre for Health Economics, Swansea, Wales, UK; College of Human and Health Sciences, Swansea, Wales, UK

**Author notes:** (Corresponding author) Emma Davies.

**Keywords:** Opioid Analgesics, drug prescribing, socioeconomic status, trends, pain management

## Abstract

**Objectives:** To use a proxy-measure of oral morphine equivalent dose (OMED) to determine trends in opioid burden in people with non-cancer pain and explore differences related to deprivation status.

**Design, setting and participants:** Retrospective cohort study using cross-sectional and longitudinal trend analyses of opioid prescribing data from 78% of Welsh Primary Care General Practices, whose data is shared with the Secure Anonymised Information Linkage (SAIL) databank. Anonymised data for the period 2005 to 2015, for people aged 18 or over, without a recorded cancer diagnosis and who received at least one prescription for an opioid medicine was included.

**Primary and Secondary outcomes:** A proxy-measure of oral morphine equivalence dose (OMED) was used to describe trends in opioid burden over the study period. OMED burden was stratified by 8 drug groups and deprivation, based on the quintile measures of the Welsh Index of Multiple Deprivation 2011 (WIMD2011).

**Results:** In the 11 years examined, 22 641 424 prescriptions for opioids were issued from 345 primary care general practices in Wales. Daily OMED per 1000 population increased by 94.7% (from 16 266 mg to 31 665 mg). Twenty-eight percent of opioid prescribing occurred in the most deprived quintile. More than 100 000 000mg more OMED was prescribed in the most deprived areas of Wales, compared to the least deprived. Codeine prescribing accounted for 35% of the OMED burden in Wales over the study period.

**Conclusions:** Whilst opioid prescription numbers increased 44% between 2005 and 2015, the OMED burden nearly doubled, with a disproportionate OMED load in the most deprived communities in Wales. Using OMED provides an insightful representation of opioid burden, more so than prescription numbers alone. Socio-economic differences are likely to affect pain presentation, access to support services and increase the likelihood of receiving an opioid prescription.

**Strengths and limitations of this study:**

- This study forms part of the first large-scale examination of opioid prescribing in Wales and is the first to use oral morphine equivalent dose as an outcome measure.
- Access to anonymously linked data allows more detailed examination of demographic influences on opioid prescribing.
- The study used a proxy-measure for oral morphine equivalent dose due to unavailability of anonymously linked prescription dispensing data.
- Disproportionate levels of prescribing in particular populations have been reported in many countries; further research should seek to understand the reasons for the differences and develop means to address any inequality noted.

## Introduction

The number of prescriptions for opioid medicines issued in the United Kingdom (UK) has increased substantially over the last twenty years (1-6). In particular, prescriptions for ‘strong’ opioids, such as morphine, oxycodone and fentanyl, have seen a higher increase than those classed as ‘weak’, like codeine and dihydrocodeine (1,2,6). Increases in prescribing persist, even when evidence to support using these medicines for people living with pain that is not acute, cancer or end-of-life related, is largely missing(7-11).

National and international concerns have focussed on the rising use of strong opioids (12-14). However, dose and duration of use are more likely indicators of harm or potential for dependence than the drug type (11, 15-20). The literature suggests adverse events occurred in 78% of cases for opioid use over extended periods of time (11,21,22). Similarly, increasing doses (8,15,23-25) have been associated with depression and anxiety (16,26,27) and an increased risk of dependence and misuse (20,28-30).

This is a particular concern in areas of high socioeconomic deprivation, where prescribing of potentially dependence forming medicines, including opioids, has been found to be more prevalent, especially for chronic, non-cancer pain in the UK (2,4,6,19,31-34) and internationally (35). Deprivation is a predictor for poor health outcomes and higher incidence of mental health disorders (36), and concomitant use of other medicines, such as benzodiazepine and anti-depressants, and opioids confer additional risk of harm to the user (19,34,37).

Wales has historically high levels of deprivation (38). In 2018, 23% of the Welsh population lived in poverty, which is more than in England, Scotland or Northern Ireland (39). Swansea, Rhondda Cynon Taff and Neath Port Talbot, some of the most deprived areas in Wales (40), have the highest opioid-related death rates in England and Wales (41) and higher numbers of non-fatal opioid overdoses were reported in Wales compared to England (42). However, a comprehensive analysis of Wales-wide opioid prescribing has only recently been undertaken (4).

The aim of this study was to examine the changes in opioid prescribing in Wales between 2005 and 2015. A proxy-measure of oral morphine equivalent dose (OMED) was used to standardise data and allow comparisons of the opioid burden from each drug in use. Data was stratified by deprivation quintile, to determine if opioid burden varies depending on socio-economic differences.

## Method

This research was approved by the Information Governance Review Panel (IGRP) of the Secure Anonymised Information Linkage databank (SAIL), based in Swansea University (SAIL identification number: 0507).

### Data source

The study used anonymised data held in the Secure Anonymised Information Linkage(SAIL) system, which is part of the national e-health records research infrastructure for Wales (43,44).

The main data source was the Primary Care General Practice (PCGP) dataset, which contained complete data from 1 January 2005 until 31 December 2015. A new dataset was formed using one line per opioid prescription, linked to individual patient demographic data using unique Anonymised Linkage Fields (ALFs). The ALF allows cross-linking between different existing datasets, providing a record of all healthcare interactions for any individual whose data is available to SAIL. Primary Care General Practice data was linked to the Welsh Index of Multiple Deprivation 2011 (WIMD2011), based on the Local Super Output Areas (LSOAs) listed on the PCGP records.

### Opioid prescriptions

Prescriptions are automatically assigned read codes at issue as a consistent means to identify data of interest (1,3,43,44). Read codes are a thesaurus of clinical terms used to record all interactions, diagnoses and interventions in Primary Care settings in Wales. A list of read codes was compiled for all prescribable oral and transdermal opioid medicines used for analgesia, including combination products, e.g. paracetamol and codeine (co-codamol), using the NHS Information Authority’s Clinical Terminology Browser and accessed via the SAIL secure gateway. Products licensed for the management of misuse and injectable opioids, which are reserved for palliative care, were excluded.

Only data for people aged 18 years or older between 2005 and 2015 without a recorded cancer diagnosis (identified using read codes for cancer diagnoses or treatment) at any time between 2004 and 2015 were included in the analysis.

All data were subjected to repeated cross-sectional sampling to determine prescribing trends over the study period.

### Oral morphine equivalent dose

Oral morphine equivalent dose (OMED) is normally calculated by multiplying the dose for each prescription (or a combination of doses, e.g. 10mg + 30mg tablets to give a total of 40mg per dose), by the equi-analgesic ratio of the opioid in question (1). The number of days’ supply provided by each prescription is then divided by the numerical daily dose (NDD) taken from the free text on the prescription (e.g. ‘one tablet to be taken twice daily’). This way, an OMED can be calculated per prescription or per individual over longer time periods (e.g. annual OMED) (1,2).

However, at the time of this study, dispensing data (42) could not be anonymously linked into SAIL datasets. Whilst the prescribed product, including strength, was recorded, details such as directions or quantity prescribed was unavailable to calculate the prescribed oral morphine equivalent dose (OMED) for each individual.

Therefore, each opioid product was allocated its OMED value based on conversion tables available for clinical practice (8,43) and dose and then multiplied by the recommended dose per day, as available from the British National Formulary (46) or the summary of product characteristics (47). The OMED for each product was multiplied by the number of prescriptions issued each year to determine the annual totals. Results were stratified by drug, with less frequently used medicines grouped as ‘other’ opioids. This included diamorphine (oral preparations), dipipanone, hydromorphone, meptazinol, methadone (tablets), pentazocine, pethidine and tapentadol.

A secondary analysis used proxy-OMED totals for products which had a daily dose of 120mg OMED or more, classed as ‘high dose’ opioid prescribing (8).

### Deprivation scores

The Welsh Index of Multiple Deprivation (WIMD) is the official measure used by Welsh Government to determine relative deprivation of areas within Wales (40). The WIMD is a weighted total score of deprivation based on income (23.5%), employment (23.5%), health (14%), education (14%), geographical access of services (10%), community safety (5%), physical environment (5%) and housing (5%). Scores are not linear, so areas in group 2 are not twice as deprived as those in group 4. Indices are published every 3 years. The 2011 Index was used for this study. Data are presented in quintiles, with WIMD1 being the most deprived areas and WIMD5 the least deprived.

### Measuring utilisation

The number of prescriptions and number of patients per year, per drug were calculated in repeat, cross-sections for each year and further stratified into drug and deprivation group. Data were standardised to annual population size using data from the Office of National Statistics (ONS) (48) and StatsWales (49). Deprivation data was adjusted by each quintile’s annual population.

### Data analysis

Data was extracted from the study tables within SAIL using Structured Query Language-code searches in Eclipse software. Trends were reported standardised per 1000 population. Percentage change rate of number of prescriptions issued and number of people receiving prescriptions over the study period were also noted.

Data was stratified into 8 groups. The 7 most commonly prescribed drugs were buprenorphine, codeine, dihydrocodeine, fentanyl, morphine, oxycodone and tramadol. The ‘other’ group included dextropropoxyphene, meptazinol, pethidine and tapentadol. Statistical analysis was conducted using SPSS 25 software and figures drawn using Excel 16.30.

### Patient and public involvement statement

There was no direct patient involvement in development and design of this study. However, the SAIL databank has members of the public who provide advice and give recommendations on safeguarding and ethical approval via a Consumer Panel. Panel members also provide input to the IGRP, which approves all data applications.

## Results

Prescribing data were extracted from 345 of the 443 (78%) Primary Care General Practices in Wales. A total of 22 641 424 prescriptions for opioids were included in this analysis. Between 2005 and 2015, the number of opioid prescriptions increased by 44% from 692 to 994 prescriptions per 1000 population annually. The total daily OMED, issued in all included practices in Wales doubled in the 11 years examined, rising from 37 662 651 mg to 76 428 768 mg. When adjusted to population, daily OMED per 1000 population increased by 94.7% (from 16 266 mg to 31 665 mg) over the study period **(Table 1)**.

**Table 1:** Daily oral morphine equivalent dose (mg) issued on prescription, given as annual totals and adjusted to population, stratified by drug

|  |  | Oral or transdermal opioids |  |  | Products with a daily dose >120mg OMED |  |  |  |
| --- | --- | --- | --- | --- | --- | --- | --- | --- |
|  |  | Annual total daily oral morphine equivalent (mg) dose prescribed | Annual total daily oral morphine equivalent dose (mg) per 1000 population | Oral morphine equivalent dose per prescription issued (mg) | Annual number of prescriptions issued per 1000 population | Annual total daily oral morphine equivalent dose (mg) | Annual total daily oral morphine equivalent dose (mg) per 1000 population | Annual total daily oral morphine equivalent dose (mg) per prescription |
| Buprenorphine | 2005 | 977 464 | 422 | 98 | 4 | 474 808 | 205 | 136 |
|  | 2015 | 2 756 458 | 1 142 | 37 | 31 | 817 904 | 339 | 139 |
|  | Percentage/rate change (%) | 182.0 | 170.5 | -61.7 | 606.3 | 72.3 | 65.3 | 2.5 |
| Codeine | 2005 | 13 743 115 | 5 916 | 17 | 357 | - | - | - |
|  | 2015 | 25 593 382 | 10 581 | 19 | 549 | - | - | - |
|  | Percentage/rate change (%) | 86.2 | 78.8 | 16.2 | 53.9 | - | - | - |
| Dihydrocodeine | 2005 | 4 368 806 | 1 887 | 12 | 154 | - | - | - |
|  | 2015 | 3 471 460 | 1 438 | 13 | 109 | - | - | - |
|  | Percentage/rate change (%) | -20.5 | -23.8 | 7.7 | -29.2 | - | - | - |
| Fentanyl | 2005 | 2 695 290 | 1 164 | 186 | 6 | 2 173 920 | 939 | 244 |
|  | 2015 | 6 496 270 | 2 691 | 147 | 18 | 4 712 410 | 1 952 | 259 |
|  | Percentage/rate change (%) | 141.0 | 131.2 | -21.2 | 193.2 | 116.8 | 108.0 | 6.5 |
| Morphine | 2005 | 3 293 220 | 1 422 | 86 | 17 | 2 554 920 | 1 103 | 135 |
|  | 2015 | 17 047 800 | 7 063 | 68 | 104 | 11 572 760 | 4 795 | 127 |
|  | Percentage/rate change (%) | 417.7 | 396.6 | -20.6 | 525.6 | 353.0 | 334.5 | -5.5 |
| Oxycodone | 2005 | 1 316 480 | 569 | 105 | 5 | 872 240 | 377 | 179 |
|  | 2015 | 6 165 400 | 2 554 | 100 | 26 | 4 003 400 | 1 659 | 184 |
|  | Percentage/rate change (%) | 368.3 | 349.3 | -4.9 | 372.4 | 359.0 | 340.3 | 2.8 |
| Tramadol | 2005 | 7 865 695 | 3 397 | 36 | 95 | - | - | - |
|  | 2015 | 14 252 335 | 5 905 | 38 | 156 | - | - | - |
|  | Percentage/rate change (%) | 81.2 | 73.8 | 5.7 | 64.4 | - | - | - |
| Other | 2005 | 3 446 735 | 1 719 | 27 | 56 | 2 0400 | 9 | 47 |
|  | 2015 | 699 711 | 347 | 58 | 5 | 339 700 | 141 | 137 |
|  | Percentage/rate change (%) | -79.7 | -79.8 | 117.4 | -91.0 | 1 565.2 | 1 497.5 | 190.6 |

Codeine was the most commonly prescribed opioid, with just under 12.5 million prescriptions issued and the highest annual total OMED prescribed for the study duration **(Figure 1)**. Its OMED per 1000 population increased by 79%, from 5916 mg to 10 581 mg. Tramadol was the second most commonly prescribed opioid in Wales. Its use increased by 74%, from 3397 mg to 5905 mg OMED per 1000 population, although annual total OMED reduced from 2014 **(Figure 1)**.

**Figure 1:**
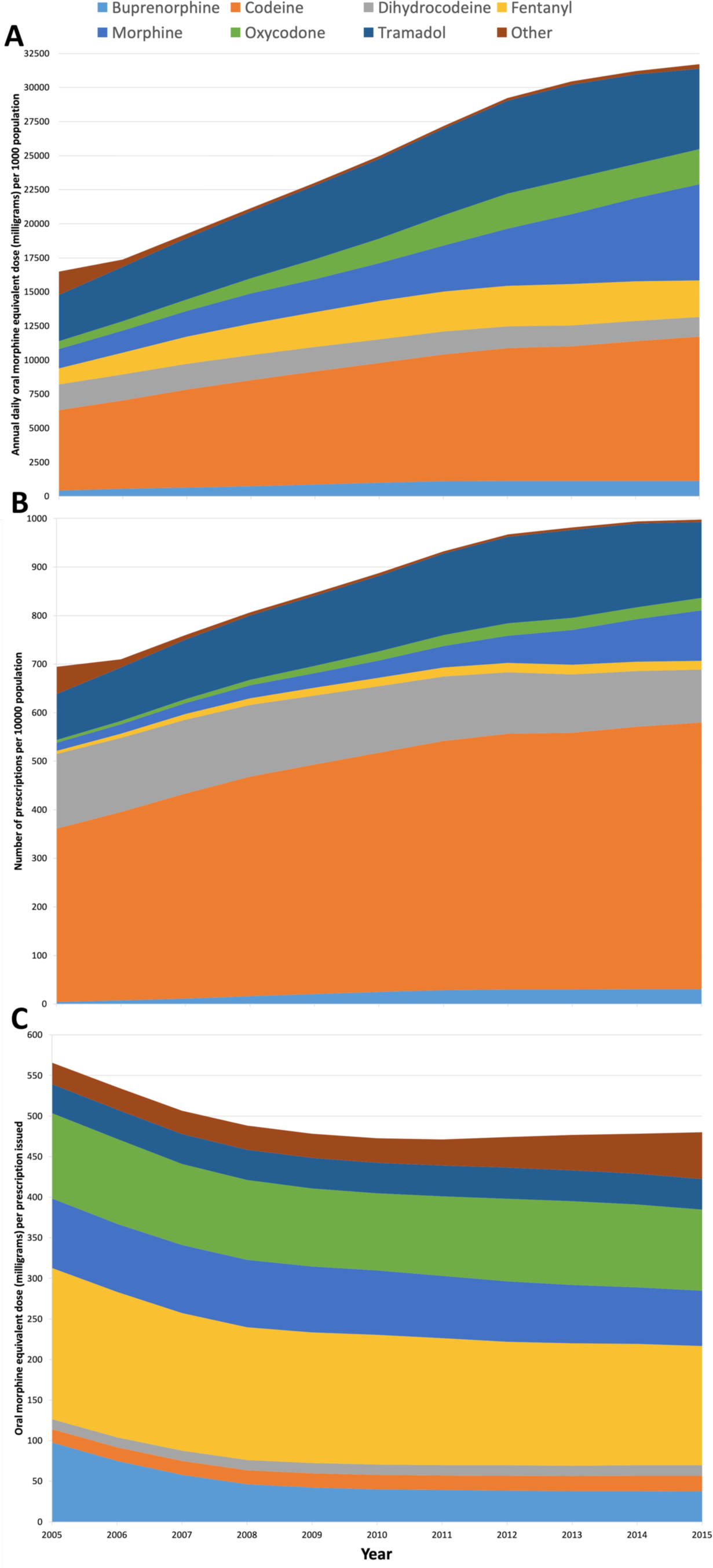
Trends in opioid prescribing across Wales, 2005 - 2015. (A) Annual daily oral morphine equivalent (OMED) in mg per 1000 population, stratified by drug. (B) Number of prescriptions per 1000 population stratified by drug (C) Annual daily OMED (mg) per prescription issued and stratified by drug

Large increases were noted in ‘strong’ opioids (morphine, oxycodone, fentanyl and buprenorphine) during the study **(Figure 1)**. Morphine OMED increased by 397%, from 1422mg to 7063mg per 1000 population. By 2015, morphine was prescribed around 3 times the equivalent dose of either oxycodone (increased 349%, from 569mg to 2554mg per 1000 population) or fentanyl (increased 131%, from 1164mg to 2 691mg per 1000 population). Morphine OMED was around 6 times that of buprenorphine, which itself increased 171%, from 422 to 1142mg per 1000 population.

Overall, 71% of the total opioid burden in the areas of Wales covered by the SAIL databank was accounted for by 3 drugs; codeine (35%), tramadol (22%) and morphine (14%). Total OMED for each drug, prescribed over the 11-year study period, were statistically different to each other (p<0.001, H=73.5, η^2^ =0.8). Post-hoc analysis confirmed differences between total OMEDs of codeine compared to dihydrocodeine (p<0.001), buprenorphine (p<0.001), oxycodone (p<0.001) and the ‘other’ category of opioids (p<0.001). Tramadol OMED demonstrated significant differences when compared to buprenorphine (p<0.001), oxycodone (p<0.05) and ‘other’ opioids (p<0.001). Morphine OMED was statistically different to buprenorphine (p<0.05) and the ‘other’ category (p<0.001).

### Oral morphine equivalent dose per prescription

Between 2005 and 2015, for all opioids, OMED per prescription increased by 36%, from 23mg to 32mg. Analysis of the eight drug groups demonstrated a statistically significant difference in OMED per prescription, largely between weak and strong opioids (p<.001, H=82.4, η^2^=0.9).

Despite the large increases in the number of prescriptions issued, OMED per prescription reduced for the four main ‘strong’ opioids, morphine (-21%, from 86mg to 68mg), oxycodone (-5%, from 105mg to 100mg), fentanyl (-21%, from 186mg to 147mg) and buprenorphine (-62%, from 98mg to 37mg), **(Table 1)**. Buprenorphine OMED reduction resulted from falling use of twice-weekly, higher dose patches (>35micrograms per hour), which outweighed the higher number of prescriptions for lower dose weekly patches (5-20 micrograms per hour).

Whilst the number of prescriptions for ‘other’ opioids decreased by 91% (from 56 to 5 per 1000 population) over the study period, the OMED per prescription more than doubled from 27mg to 58mg per prescription. Prescription numbers notably increased from 2011 onwards, when tapentadol was released onto the UK market. Codeine, dihydrocodeine and tramadol dose per prescription increased over the 11 year period.

### High dose opioid prescribing

Based on the strength of preparation prescribed, five groups of drugs demonstrated dosing of 120mg OMED and over between 2005 and 2015 **(Table 1)**. When compared to the total OMED prescribed in the study period, statistically significant differences were established between them (p=0.000, H=45.7, η^2^=0.8). High-dose fentanyl OMED was confirmed to be significantly larger than OMED for buprenorphine (p<0.001), morphine (p<0.001) and ‘other’ opioids (p<0.001), as was oxycodone OMED when compared to morphine (p<0.005) and ‘other’ opioids (p0<.001).

Whilst morphine (335% increase from 1103mg to 4795mg per 1000 population) and oxycodone (340% increase from 377mg to 1659mg per 1000 population) saw similar percentage increases, the total OMED for high-dose morphine was nearly three times that of oxycodone throughout the study **(Figure 2)**. The large percentage increase in ‘other’ opioids prescribed at 120mg OMED or above (1497% from 9mg to 141mg per 1000 population) was entirely due to tapentadol prescribing from 2011 onwards. Tapentadol prescribing tended to be at high OMED levels.

**Figure 2:**
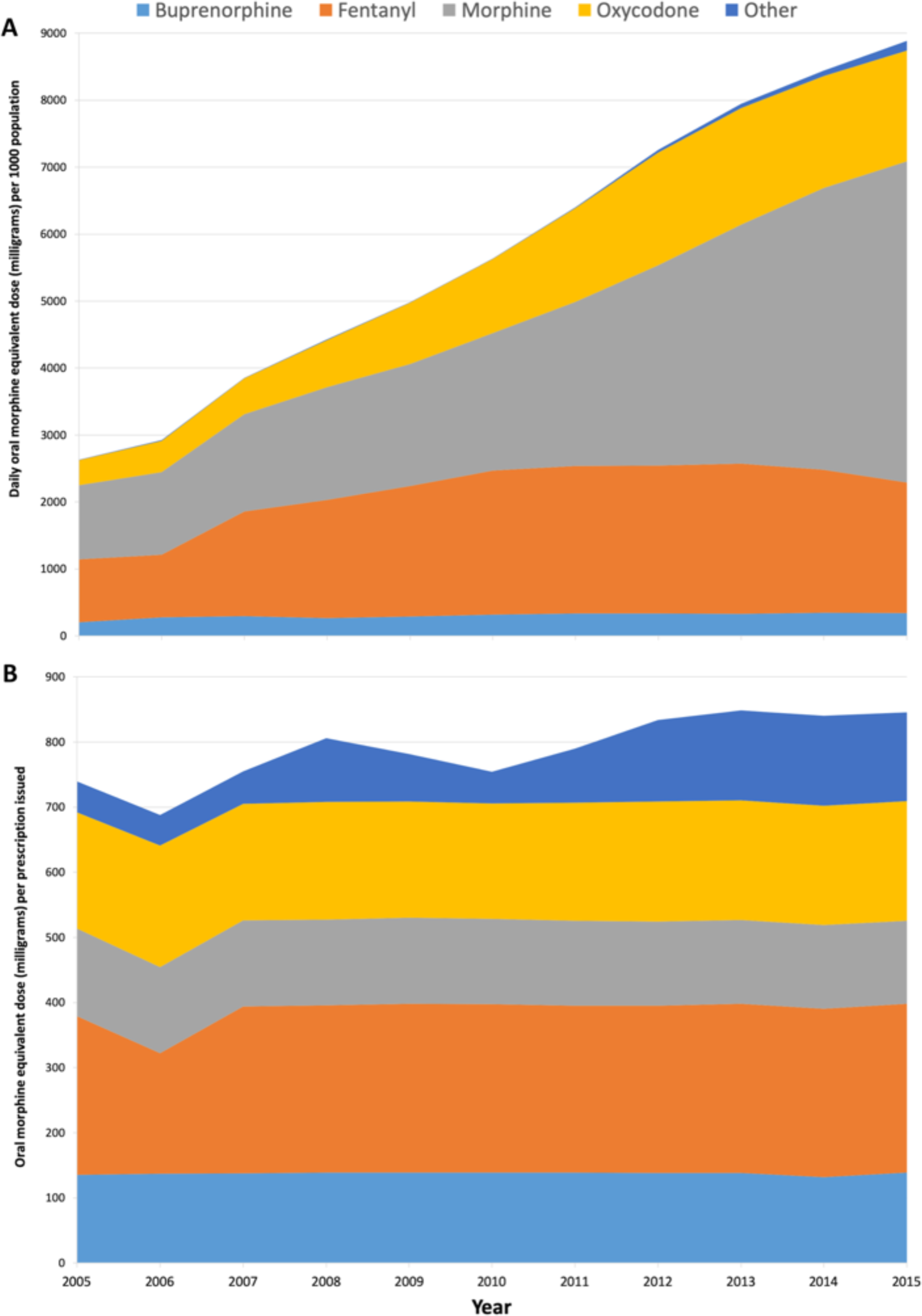
Trends in high-dose (>120mg OMED per day) opioid prescribing in Wales, 2005-2015. (A) Daily OMED (mg) per 1000 population stratified by drug (B) OMED (mg) per prescription issued stratified by drug

### Opioid prescribing trends by deprivation

The trends in annualised daily OMED of all oral and transdermal opioids stratified by the Welsh Index of Multiple Deprivation (WIMD2011) are illustrated in Figure 3. Over the 11 years of the study, the most deprived areas (WIMD1) examined were prescribed 100 711 696mg more OMEDs than the least deprived (WIMD5) **(Table 2)**.

**Table 2:** Trends in oral morphine equivalent prescribing stratified by deprivation (Welsh Index of Multiple Deprivation WIMD2011). WIMD1=most deprived, WIMD5=least deprived

|  |  | Oral or transdermal opioids |  |  | Products with a daily >120mg OMED |  |  |  |
| --- | --- | --- | --- | --- | --- | --- | --- | --- |
|  |  | Oral morphine<br>equivalent dose<br>(mg) prescribed | Oral morphine<br>equivalent dose<br>(mg) per 1000<br>population | Oral morphine<br>equivalent dose<br>(mg) per<br>prescription | Oral morphine<br>equivalent dose<br>(mg) per day | Total oral<br>morphine<br>equivalent dose<br>(mg) per 1000<br>population | Oral morphine<br>equivalent dose<br>(mg) per<br>prescription | Oral morphine<br>equivalent dose<br>(mg) per person<br>issued<br>prescriptions |
| WIMD1 |  |  |  |  |  |  |  |  |
|  | 2005 | 10 319 636 | 21 757 | 22 | 1 466 652 | 3092 | 166 | 822 |
|  | 2015 | 21 167 919 | 43 176 | 31 | 5 674 237 | 11574 | 152 | 1111 |
|  | Percentage/rate change<br>(%) | 105.1 | 98.4 | 41.0 | 286.9 | 274.3 | -7.8 | 35.2 |
|  | Total prescribed | 176 824 265 |  |  | 37 943 319 |  |  |  |
| WIMD2 |  |  |  |  |  |  |  |  |
|  | 2005 | 8 590 375 | 18 203 | 24 | 1 208 800 | 2561 | 162 | 834 |
|  | 2015 | 17 399026 | 35 475 | 32 | 4 837 285 | 9863 | 158 | 1078 |
|  | Percentage/rate change<br>(%) | 102.5 | 94.9 | 36.3 | 300.2 | 285.0 | -2.5 | 29.3 |
|  | Total prescribed | 146 459 878 |  |  | 32 622 635 |  |  |  |
| WIMD3 |  |  |  |  |  |  |  |  |
|  | 2005 | 7 684 060 | 17 108 | 24 | 1 373 446 | 3058 | 175 | 847 |
|  | 2015 | 15 342 942 | 32 564 | 33 | 4 642 419 | 9853 | 155 | 1010 |
|  | Percentage/rate change<br>(%) | 99.7 | 90.3 | 33.1 | 238.0 | 222.2 | -11.4 | 19.2 |
|  | Total prescribed | 129 880 669 |  |  | 33 344 376 |  |  |  |
| WIMD4 |  |  |  |  |  |  |  |  |
|  | 2005 | 5 374 595 | 12 242 | 25 | 1 104 120 | 2515 | 188 | 911 |
|  | 2015 | 10 878 897 | 23 534 | 33 | 3 314 817 | 7171 | 157 | 988 |
|  | Percentage/rate change<br>(%) | 102.4 | 92.2 | 28.9 | 200.2 | 185.1 | -16.5 | 8.5 |
|  | Total prescribed | 93 691 687 |  |  | 25 420 454 |  |  |  |
| WIMD5 |  |  |  |  |  |  |  |  |
|  | 2005 | 4 486 035 | 9 381 | 24 | 697 496 | 1459 | 163 | 662 |
|  | 2015 | 8 721 170 | 17 557 | 30 | 2 228 629 | 4487 | 154 | 829 |
|  | Percentage/rate change<br>(%) | 94.4 | 87.2 | 27.9 | 219.5 | 207.6 | -5.5 | 25.2 |
|  | Total prescribed | 76 112 569 |  |  | 16 958 635 |  |  |  |

**Figure 3:**
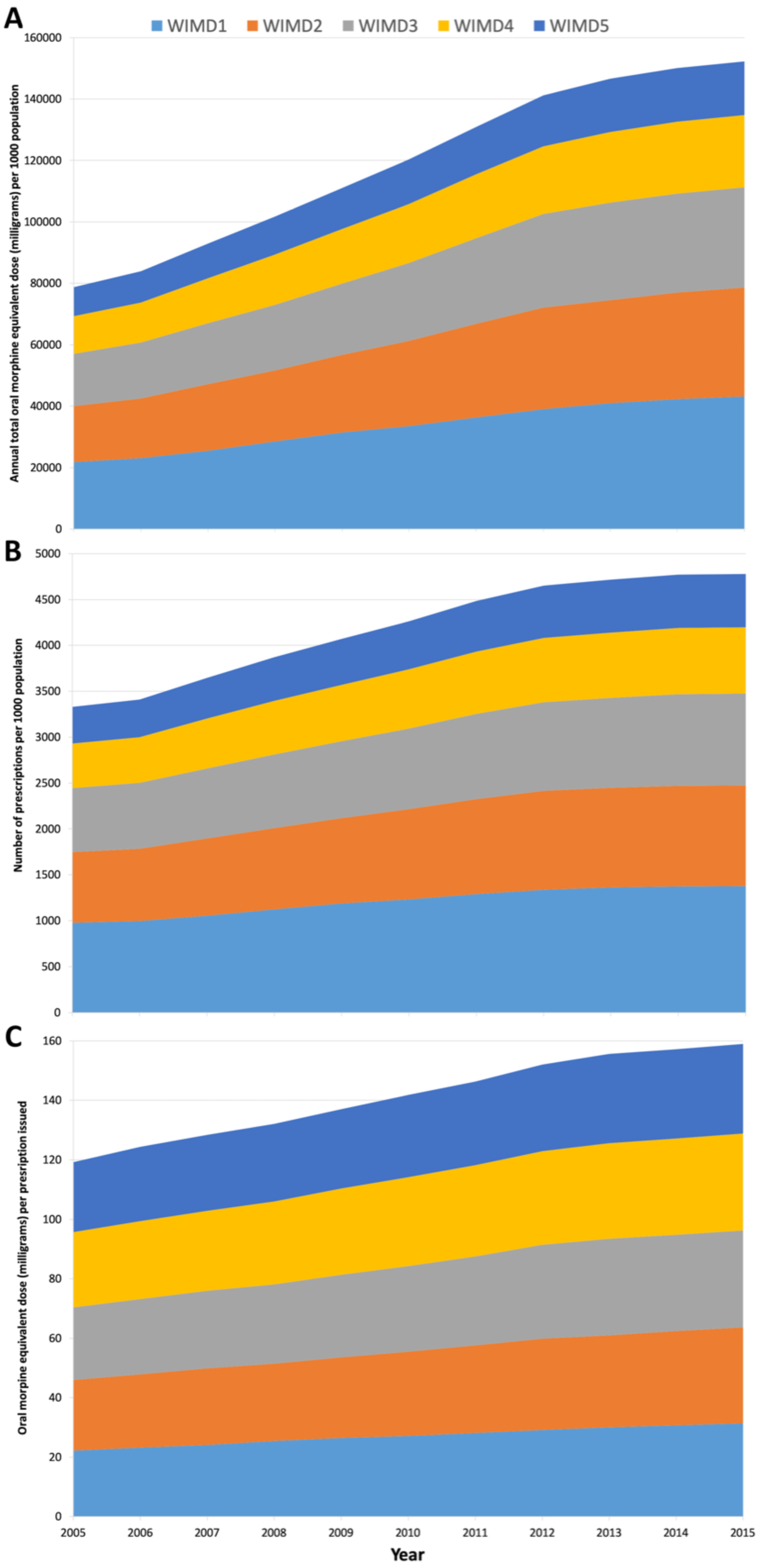
Trends in opioid prescribing across Wales, 2005 - 2015. (A) Annual daily oral morphine equivalent (OMED) (mg) per 1000 population, stratified by deprivation. (B) Number of prescriptions per 1000 population stratified by deprivation (C) Annual daily OMED (mg) per prescription issued and stratified by deprivation. Welsh Index of Multiple Deprivation 2011 (WIMD2011) where WIMD1=most deprived, WIMD5=least deprived

Between 2005 and 2015, OMED doubled in all but the least deprived (WIMD5) areas **(Table 2)**, with 28% (176 824 265mg of 622 969 068mg) of the total OMED prescribed issued in the most deprived areas of Wales. In contrast, 12% (76 112 569mg) were prescribed in the least deprived areas. Throughout the study, the OMED prescribed in WIMD1 areas remained more than twice those noted in WIMD5 areas **(Table 2)** for both total OMED (mg) and OMED per 1000 population. Despite large percentage increases in all quintiles, the total OMED prescribed in each area was significantly different (p<0.001, H=34.5, η^2^=0.61). The least deprived areas had significantly lower prescribed OMED than quintiles 1 to 3 (WIMD1 p<0.001, WIMD2 p<0.001 and WIMD3 p<0.05). The total OMED prescribed in the most deprived areas (WIMD1) was significantly higher compared to that in WIMD4 areas (p<0.05).

### OMED per prescription

OMEDs per prescription were similar in all quintiles, despite the large differences in the actual number of prescriptions issued **(Figure 3)**. The percentage change in OMED per prescription increased with greater deprivation, with WIMD1 areas seeing increases of 41%, from 22mg to 31mg per prescription, whereas WIMD5 areas increased by 28%, from 24mg to 30mg per prescription between 2005 and 2015. No statistically significant difference in the total OMED per prescription between deprivation quintiles was found (p>0.05, H=5.3, η^2^=0.03).

### OMED per person

Oral morphine equivalent dose per person was significantly different between quintiles (p<0.05, H=11.1, η^2^=0.6) **(Table 2)**. The OMED per person was confirmed to be significantly greater in the two most deprived areas (WIMD1 p<0.05, WIMD2 p<0.05) compared to the least deprived quintile (WIMD5). In WIMD1 areas, a 79% increase led the dose per person to rise from 96mg to 173mg in 11 years. Smaller increases were seen in WIMD2 (70%, from 99mg to 169mg per person), WIMD3 (65%, from 98mg to 161mg per person), WIMD4 (61%, from 93mg to 151mg per person). A 60% increase in WIMD5 areas, from 80mg to 128mg per person resulted in a 35% difference in OMED per prescription between the least and most deprived areas in Wales.

### High-dose opioid prescribing by deprivation

The total OMED prescribed for products with a daily OMED of 120mg and over tripled in each deprivation quintile between 2005 and 2015 **(Table 2)**. As observed with all opioid prescribing, the most deprived areas had the greatest high-dose OMED prescribed. The total of 37 943 319mg prescribed over 11 years was 2.24 times more than in the least deprived quintile, where 16 958 635mg were prescribed in the same period. The differences between high-dose OMED in each quintile were found to be statistically significant with small effect size (p=0.002, H=17.1, η^2^=0.26). The total high-dose OMED in the WIMD5 areas was significantly different to those in WIMD1 (p<0.05), WIMD2 (p<0.05) and WIMD3 (p<0.05) areas.

After adjustment for population size per quintile, the large differences in OMED per 1000 population dependent on the level of deprivation were maintained with significant differences, although small effect size (p=0.001, H18.5, η^2^=0.29). OMED per 1000 population was significantly lower in the least deprived areas of Wales compared to quintiles 1 to3, all with p<0.05. In 2015, around 17% more high-dose OMED was prescribed in WIMD1 areas (11574mg per 1000 population) than in the next most deprived WIMD2 (9863mg per 1000 population). Even though high-dose OMED between 2005 and 2015 tripled in WIMD5 areas, OMED prescribed was still 156% higher in WIMD1 areas at the end of the study **(Figure 4)**.

**Figure 4:**
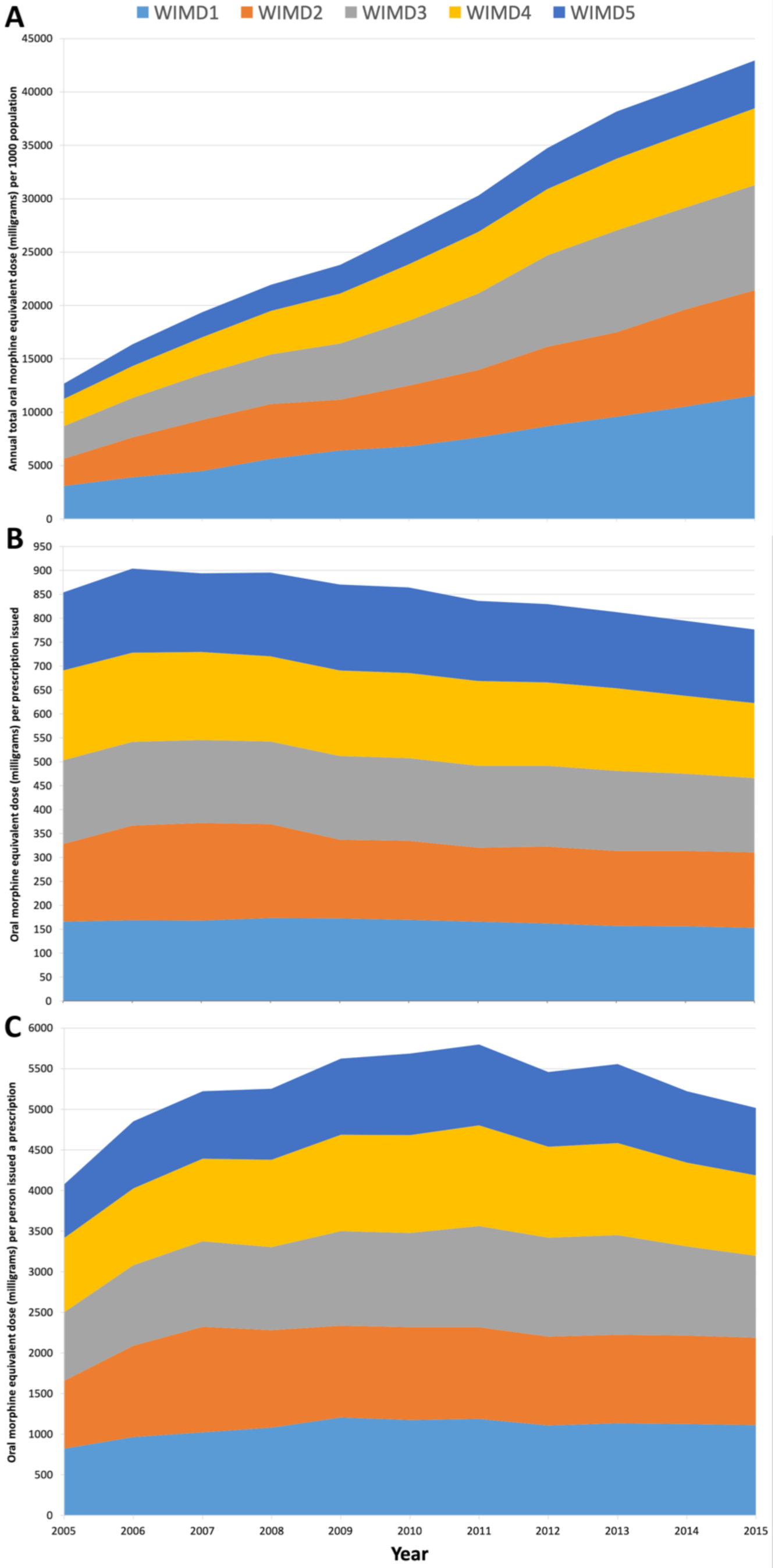
Trends in high-dose opioid (>120mg OMED) prescribing across Wales, 2005 - 2015. (A) Annual daily oral morphine equivalent (OMED) in mg per 1000 population, stratified by deprivation. (B) OMED (mg) per prescription stratified by deprivation (C) OMED (mg) per person issued a prescription and stratified by deprivation. Welsh Index of Multiple Deprivation 2011 (WIMD2011) where WIMD1=most deprived, WIMD5=least deprived

## Discussion

The results of this retrospective database study demonstrate a marked increase in opioid burden in Wales between 2005 and 2015. The rate of increase slowed toward the end of the study period. An increase of 44% in the number of opioid prescriptions per 1000 population between 2005 and 2015 in Wales has been reported (4). Using the proxy measure of OMED described, the opioid burden on the population over the study period nearly doubled.

The data demonstrates the disproportionate opioid burden felt by people living in the most deprived areas of Wales. Increasing deprivation was linked to the receipt of higher doses, higher OMED and a higher opioid burden per person. Whilst rises in percentage terms were similar in all WIMD2011 quintiles, significant differences were consistent between the most and least deprived areas throughout the 11 years examined without evidence that the trend was beginning to change.

Despite a low OMED per prescription, codeine is used widely throughout Wales and places a major burden on the population. Morphine was the most burdensome strong opioid, although both oxycodone and fentanyl had higher OMED (mg) per prescription issued.

This study identified rising trends in opioid prescribing in Wales, similar to those previously reported in other parts of the UK (1,3,6,32,33,50).

Examining trends by prescription numbers alone is likely to underestimate the opioid burden within a population. Curtis et al demonstrated a 34% growth in prescription numbers resulted in a 127% increase in OMED burden (6). Similarly, as shown in this study, a 44% increase in prescription numbers in Wales translated into a 95% increase in opioid burden using the proxy-OMED measure described.

Codeine is referenced as a ‘weak’ opioid (8,46) and its risks may therefore be downplayed. However, deaths in Wales associated with codeine, including as part of compound preparations (e.g. co-codamol), increased from 12 in 2005 to 16 in 2015 (51). Furthermore, following the end of the study period, codeine-associated deaths increased a further 50%, with 24 reported fatal incidents in 2018, making codeine responsible for 15% of opioid associated deaths in Wales in 2018 (n=164) (51). Since codeine is also available to purchase over-the-counter (OTC), prescribing data is an underestimate of population exposure to the drug (31, 52)

Higher OMED per prescription for non-morphine strong opioids may point to prescribers misunderstanding oral morphine equivalence (53-55) and using higher doses of non-morphine opioids than are recommended (8). Tapentadol was responsible for a rapid increase in OMED for ‘other’ opioids since its introduction to the UK market in 2011. This has been noted in England as well (6). By the end of the study period, high dose tapentadol OMED per prescription was greater than that of morphine.

This study has demonstrated substantial increases in opioid prescribing, with higher levels in more deprived populations, which was also previously reported in England (2,33) and internationally (56-59. Furthermore, much of the focus of the opioid crisis in the US has been on rural, deprived communities where access to, and affordability of, healthcare services and social support are poor (35,56,60).

Increased levels of prescribing in areas of high socio-economic deprivation have been linked to greater reported pain intensity (32). However, limited evidence supports the notion that opioids are effective at reducing pain, particularly in the longer term (8,18,61). High-dose opioids (above 120mg OMED) have also been associated with increased levels of pain (62,63). In the context of this and previous studies (2,6,32,33, the implications of the increased prescribing of opioids in more deprived areas are concerning as it exposes the most vulnerable people to higher levels of medicines that may be ineffective at best and may even cause additional health and well-being complications (11).

This study is the first to utilise a large dataset including routine data from 78% of Primary Care practices in Wales to map opioid prescribing trends (4). Large sets of prescribing and diagnostic data have been validated as an accurate means for conducting healthcare population research (64,65) as they reduce recall bias and regional variation. Moreover, any person registered with included practices and prescribed an opioid medicine was included in the analysis, thus avoiding selection bias. Furthermore, this is the first study conducted using Welsh data that utilised an OMED measure to better understand the burden of opioid prescribing on the population. Using the linkage systems within SAIL datasets, data from people with a recorded cancer diagnosis, could be excluded from analysis. While recent studies of opioid prescribing in the UK based on large datasets merely assumed that the majority of prescribing was attributable to persisting, non-cancer pain based on the longevity of prescribing and the dose forms used (2,6,50), this data confidently reflects prescribing for non-cancer pain.

However, prescribing data alone provides an indication of intention to treat but does not confirm consumption. It also does not give an indication of the diagnosis or how long an individual might have been using the medication. Moreover, the data presented here did not identify people receiving more than one opioid medicine and who would have a higher individual OMED burden.

It was not possible to access linked dispensing data, which would provide the detail required to more accurately calculate OMED. This resulted in the development of a proxy measure which provides a daily estimation of OMED, multiplied by the number of prescriptions as described above, and required a number of assumptions to be made. Without knowing prescription duration, i.e. how many daily doses were provided per prescription, the annualised OMEDs presented here may be an underestimation of the opioid burden. However, the trends described are similar to those reported elsewhere in the UK (1-3,6,33,50).

For this study, opioid medicines were identified by read codes and accuracy of the data extraction depended on the inclusivity of the coding used. Incomplete coding lists would result in data being lost to the analysis and could lead to an under-representation of prescribing. Also, oral and transdermal medicines were selected on the likelihood of them being used for pain management rather than the management of misuse disorder. For example, high dose sublingual buprenorphine and methadone solution were excluded. However, in rare cases, these may be prescribed as analgesia while other included opioid preparations could be used as part of misuse management. Again, this could result in an over or underestimation of prescribing. However, similar rationales for deciding which opioid products to include in analysis of Primary Care prescribing was adopted by other UK-based authors (1,3,6,33).

Using a measure of opioid burden, such as oral morphine equivalent dose gives a more accurate measure of the impact of opioid prescribing on a population. In Wales, National Prescribing Indicators are set each year by the All Wales Medicines Strategy Group (AWMSG) (66) using average daily quantities (ADQ) as the measure. ADQ is the assumed average maintenance dose per day for a drug, used for its main indication in adults. The risks of opioids are known to increase above a daily dose of 120mg OMED (8), regardless of the indication. Consequently, using a consistent measure such as OMED would provide greater understanding of the risks imposed on the population. In addition, good practice, such as using weekly prescriptions for people trialling or tapering opioids and those at risk of misuse (8), would not skew data as it does currently.

Welsh Government recently set out plans to improve access to persistent pain management support across Wales (67). It focusses on improving access to non-pharmacological and self-management support such as pain management programmes (68,69), psychological input (70) and physiotherapy (71) which have been demonstrated to have longer-term benefits and do not carry the same risks as opioids. However, as with other guidelines in this area (72, 73), the document does not include a strategy to address health inequalities in pain management support per se (64). Given the disproportionate use of opioids in the most deprived areas, it would seem essential to tackle the root causes of those differences, if anything is to improve. More research is needed to better understand the reasons opioids are still prescribed in the face of mounting evidence of harm including current concerns around dependence and misuse (8,11,31). Furthermore, greater understanding of why people living with pain continue to use opioids in spite of poor effect or additional health concerns is required.

Supporting non-pharmacological management is essential to enable people to live well with pain (74) but can be difficult to encourage in practice (75). McCrorie and colleagues (2015) concluded that problematic opioid prescribing can result from failing to meet the self-perceived needs of people living with pain and from prescribers feeling unable to negotiate alternatives (76). This was echoed by Finestone who described opioid prescribing as a surrogate for poor access to pain management support (77).

It would seem that more targeted use of resources into Primary Care, and particularly in areas of greater deprivation, would be a prudent way of improving lives and tackling the use of opioid medicines that can cause so much harm.

## Data Availability

Dataset available from the Dryad repository, DOI: https://doi.org/10.5061/dryad.1rn8pk0pf

## Contributors

ED conceived of and designed the study, collated the read-codes used for data extraction, devised the OMED proxy-measure and coded the extracted data, undertook the data analysis, drafted and revised the manuscript. BS oversaw the study design and data analysis and critically revised the manuscript. MJ oversaw the study design and statistical analysis and critically revised the manuscript. CP and JR oversaw the study design and critically revised the manuscript. All authors read and approved the final manuscript.

## Acknowledgements

This study makes use of anonymized data generated by the Secure Anonymized Information Linkage (SAIL) system, which is part of the national <u>e-health</u>records research infrastructure for Wales. We would like to acknowledge all the data providers who make anonymized data available for research.

## Patient consent for publication

Not required

## Data sharing statement

Dataset available from the Dryad repository, DOI: https://doi.org/10.5061/dryad.1rn8pk0pf

## Patient and Public Involvement statement

This research was done without patient involvement. Patients were not invited to comment on the study design and were not consulted to develop patient relevant outcomes or interpret the results. Patients were not invited to contribute to the writing or editing of this document for readability or accuracy.

